# Extraction of Viral Nucleic Acids with Carbon Nanotubes Increases SARS-CoV-2 RT-qPCR Detection Sensitivity

**DOI:** 10.1101/2021.01.22.20224675

**Authors:** Sanghwa Jeong, Eduardo G. Grandio, Nicole Navarro, Rebecca L. Pinals, Francis Ledesma, Darwin Yang, Markita P. Landry

**Affiliations:** Department of Chemical and Biomolecular Engineering, University of California, Berkeley, California 94720, United States; Department of Chemistry, University of California, Berkeley, California 94720, United States; Innovative Genomics Institute (IGI), Berkeley, California 94720, United States; California Institute for Quantitative Biosciences, QB3, University of California, Berkeley, California 94720, United States; Chan-Zuckerberg Biohub, San Francisco, California 94158, United States

## Abstract

The global SARS-CoV-2 coronavirus pandemic has led to a surging demand for rapid and efficient viral infection diagnostic tests, generating a supply shortage in diagnostic test consumables including nucleic acid extraction kits. Here, we develop a modular method for high-yield extraction of viral single-stranded nucleic acids by using ‘capture’ ssDNA sequences attached to carbon nanotubes. Target SARS-CoV-2 viral RNA can be captured by ssDNA-nanotube constructs via hybridization and separated from the liquid phase in a single-tube system with minimal chemical reagents, for downstream quantitative reverse transcription polymerase chain reaction (RT-qPCR) detection. This nanotube-based extraction method enables 100% extraction yield of target SARS-CoV-2 RNA from phosphate buffered saline in comparison to ∼20% extraction yield when instead using a commercial silica-column kit. Notably, carbon nanotubes enable extraction of nucleic acids directly from 50% human saliva, bypassing the need for further biofluid purification and avoiding the use of DNA/RNA extraction kits. Carbon nanotube-based extraction of viral nucleic acids facilitates high-yield and high-sensitivity identification of viral nucleic acids such as the SARS-CoV-2 viral genome with reduced reliance on reagents affected by supply chain obstacles.

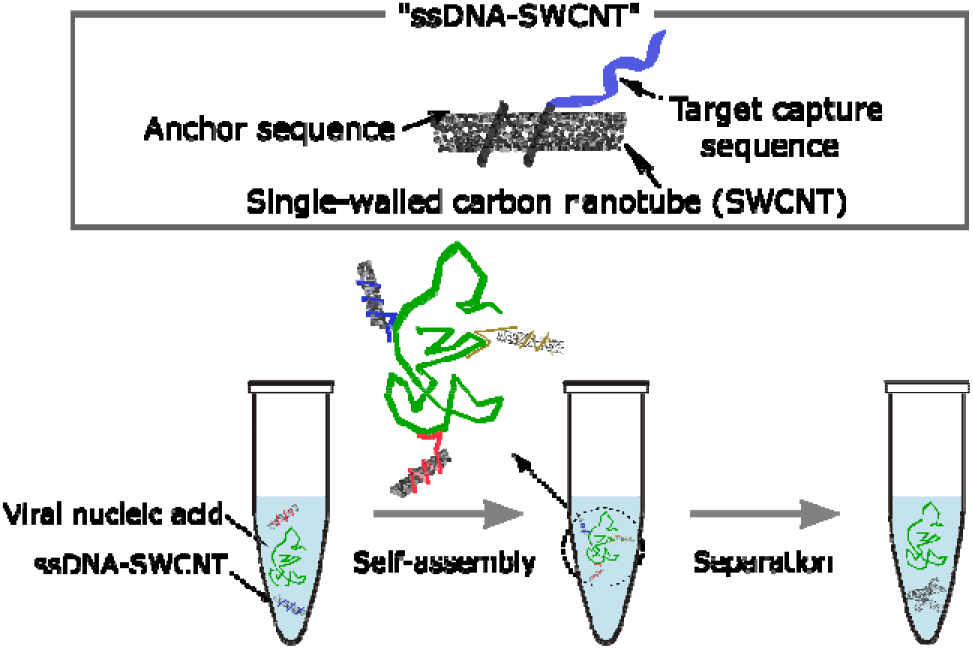

## Introduction

The emergence of the novel human coronavirus (SARS-CoV-2) in late 2019 rapidly escalated into a global pandemic. The high transmission rate and high proportion of asymptomatic infections has led to a massive, global demands for rapid and efficient viral infection diagnostic tests^1, 2^. The coronavirus pandemic has unearthed limitations of our diagnostics including insufficient supply, throughput, and variable accuracy, exacerbated by pipeline limitations in accessing reagents, specialized equipment, and trained personnel to conduct tests^2^. Low testing throughput and capacity has resulted in corresponding delays in patients receiving their testing results, an cripples contact tracing efforts^3, 4^. The two main types of diagnostic tests for virus infections, including SARS-CoV-2, are nucleic acid amplification tests (NAATs) and serological tests. NAATs are currently the gold standard, mainly using real-time reverse transcription polymerase chain reaction (RT-qPCR)^3-6^. NAATs can determine whether a person has an active infection and enables highly sensitive and quantitative diagnosis of infectious diseases. For PCR-based tests, pathogenic nucleic acid extraction from patient samples is required. However, extracting nucleic acids from a patient’s biofluid is a complicated and time-consuming task that requires trained technicians to perform, involving many processing steps^3, 4, 6^. Additionally, specialized materials such as spin columns with silica membranes or magnetic beads and chemically toxic reagents for lysis, binding, washing, and elution are needed. Together with supply chain limitations for aforementioned reagents, nucleic acid extraction constitutes a major bottleneck in current SARS-CoV-2 testing^1^. An advanced and simplified extraction methodology could increase diagnostic availability and efficiency, benefitting patient care and infection control.

Several modified protocols have been developed for extraction of nucleic acids for RT-qPCR including a chemically driven phenol/chloroform extraction method, and solid-phase extraction techniques based on nucleic acid-adsorbing substrates such as silica and cellulose^7^. For example, Wang et al. reported a DNA extraction protocol for PCR-based detection by using positively charged polyacrylamide microspheres to adsorb the negatively charged DNA and remove other patient biomolecules^8^. Kolluri et al. reported a portable nucleic acid extraction protocol from whole blood with a paper-and-plastic device for capture and purification of nucleic acids on a glass fiber membrane, followed by treatment with a chromatography paper waste pad^9^. In addition to these total nucleic acid extraction methods, affinity-based nucleic acid extraction has been also investigated by using complementary DNA or RNA sequences as capturing ligands. Selective isolation of mRNA from eukaryotic cells is commercially available with the basis of hybridization of a (dT) oligo on magnetic bead matrices to capture mRNA with an oligo (dA) tail. The magnetic beads complexed with mRNAs are next separated from total RNA and genomic DNA with application of a strong magnetic field^10^. Gopal et al. reported selective extraction of parasite *Onchocerca volvulus* DNA from the black fly by using magnetic beads with sequence-specific oligonucleotides to capture and immobilize the parasite DNA^11^. In this work, biotinylated capture oligonucleotides were bio-conjugated with streptavidin-coated magnetic beads, and this oligonucleotide based magnetic bead method improved the efficiency of pool screening of black fly vectors. In similar method, Hei et al. published the purification of gRNA of SARS-CoV with magnetic beads modified with capturing oligonucleotide^12^. While selective nucleic acid extraction with complementary DNA/RNA ligands could improve the sensitivity of downstream PCR assays for viral detection by enriching target viral nucleic acids over host DNA and RNA, aforementioned assays have not replaced or been incorporated into RT-qPCR workflows owing to their complexity, lower sensitivity, cost, or incompatibility with standard RT-qPCR protocols.

Single-walled carbon nanotube (SWCNT) is one dimensional, cylindrically shaped allotropes of carbon nanostructures with a high surface area and large aspect ratio due to their unique small diameter of □1 nm and long lateral dimension up to several micrometers^13^. Colloidal SWCNT dispersions can be prepared by the self-assembly of amphiphilic biopolymers such as nucleic acids and peptides on SWCNT surface via pi-pi stacking interactions between the aromatic bases of nucleic acids and the sp^2^ hybridized SWCNT lattice, avoiding the need for chemical conjugation of ligands to SWCNT. As such, single-stranded DNA (ssDNA) ligands can be adsorbed to SWCNT in helical configurations *via* quick few-minute probe tip sonication,^14^ avoiding time-consuming and relatively costly site-specific attachment chemistries needed to attach ssDNA to most other nanoparticles. Following non-covalent conjugation of ssDNA to SWCNT, the ssDNA on the SWCNT surface is available to hybridize and attach a target oligonucleotide with a complementary sequence, where this hybridization event can be identified with a change in the ssDNA-SWCNT fluorescence signal^15^.

Herein, we develop a high yielding method for extraction of viral single-stranded nucleic acids based on ssDNA-SWCNT constructs modified with ten unique capturing ssDNA sequences that can bind to target nucleic acids. These ssDNA-SWCNT constructs can be readily incorporated into commercial RT-qPCR testing workflows and can either increase the extraction yield of purified target nucleic acids from ∼20% (commercial kit) to ∼100% (SWCNT-based protocol), or can enable extraction of target nucleic acids in crude patient saliva biofluids, the latter of which avoids the need to purify patient samples. This viral nucleic extraction protocol permits identification of viral nucleic acids (i) with high nucleic acid extraction yield in complex biofluids, such as unprocessed saliva samples (ii) without reliance on reagents affected by supply chain obstacles, and (iii) with rapid extraction time.

## Results

To enable high-sensitivity extraction of viral nucleic acids, we implemented SWCNT as capture agents for target viral nucleic acids. SWCNTs were designed to enable (1) complexation with target viral nucleic acids via sequence-specific capture oligonucleotides on ssDNA-SWCNT constructs and (2) acid precipitation of SWCNT constructs post-capture of target viral nucleic acid (Figure 1). Ten unique ssDNA capture oligonucleotide sequences are adhered to SWCNT via (GT)_15_ anchoring segments, whereby the ten unique sequences are complementary to different segments of the SARS-CoV-2 viral RNA genome (full DNA sequences in Table 1). Each colloidal ssDNA-SWCNT construct was prepared by probe tip sonication of HiPCo SWCNT and one of the ten unique capture sequences. Of the capture sequences, nine were chosen to enable hybridization of SWCNT to the nucleocapsid (N) gene region (Target 2 – Target 10 DNA sequences), and one DNA sequence for the oligo (dA) tail (Target 1 DNA sequence). Following synthesis, these ten ssDNA-SWCNT constructs were exposed to the SARS-CoV-2 viral RNA as detailed in several experiments below, to ‘capture’ the viral nucleic acid. We hypothesize that exposing ssDNA-SWCNTs to acidic conditions would neutralize SWCNT surface charges, promoting reversible SWCNT aggregation due to attractive van der Waals interactions and reduced electrostatic repulsive forces. Therefore, the addition of hydrochloric acid (HCl) to the mixture induced precipitation of ssDNA-SWCNT constructs, to which the viral RNA is bound, enabling facile separation of bound viral RNA from the supernatant solution. The precipitates were repeatedly centrifuged and washed with DNase/RNase-free water to remove the unwanted biomolecules. Lastly, the precipitates containing ssDNA-SWCNT constructs and viral RNA were dispersed in phosphate-buffered saline (PBS) and heated at 95 °C to desorb the viral RNA from SWCNT surface. The final PBS solution containing the viral RNA can be directly used for downstream RT-qPCR for viral detection.

**Table 1.**
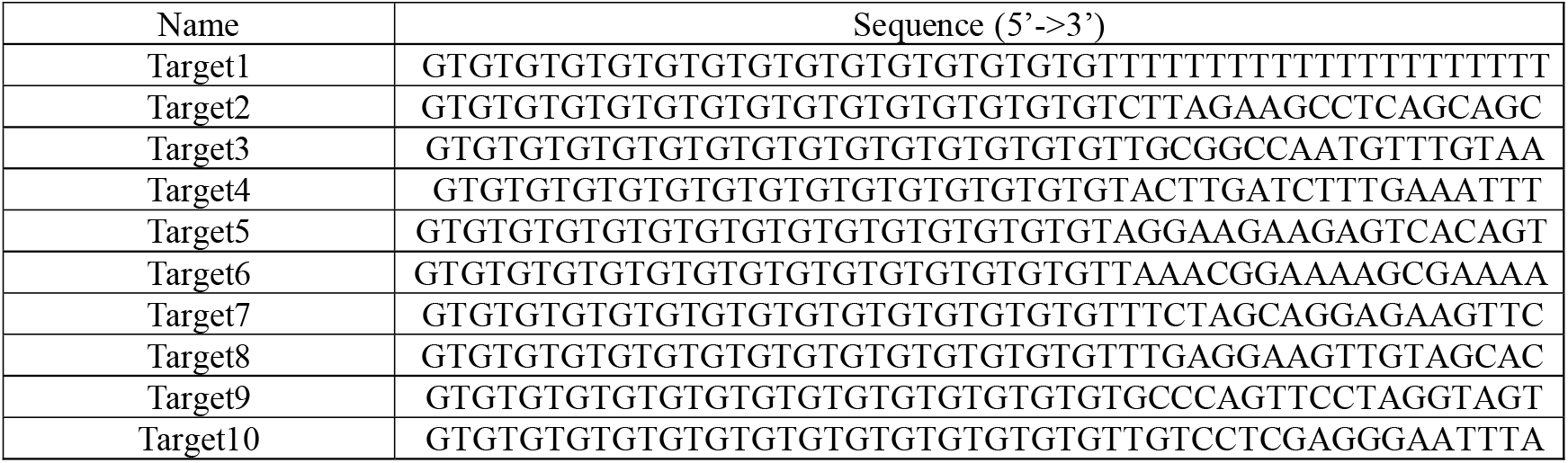
Sequences of ssDNA used for ssDNA-functionalized SWCNT constructs for SARS-CoV-2 RNA extraction. From the 5’ end, (GT)_15_ sequence is for anchoring onto the SWCNT surface, and the subsequent 18-mer sequence is for capturing SARS-CoV-2 RNA.

**Figure 1.**
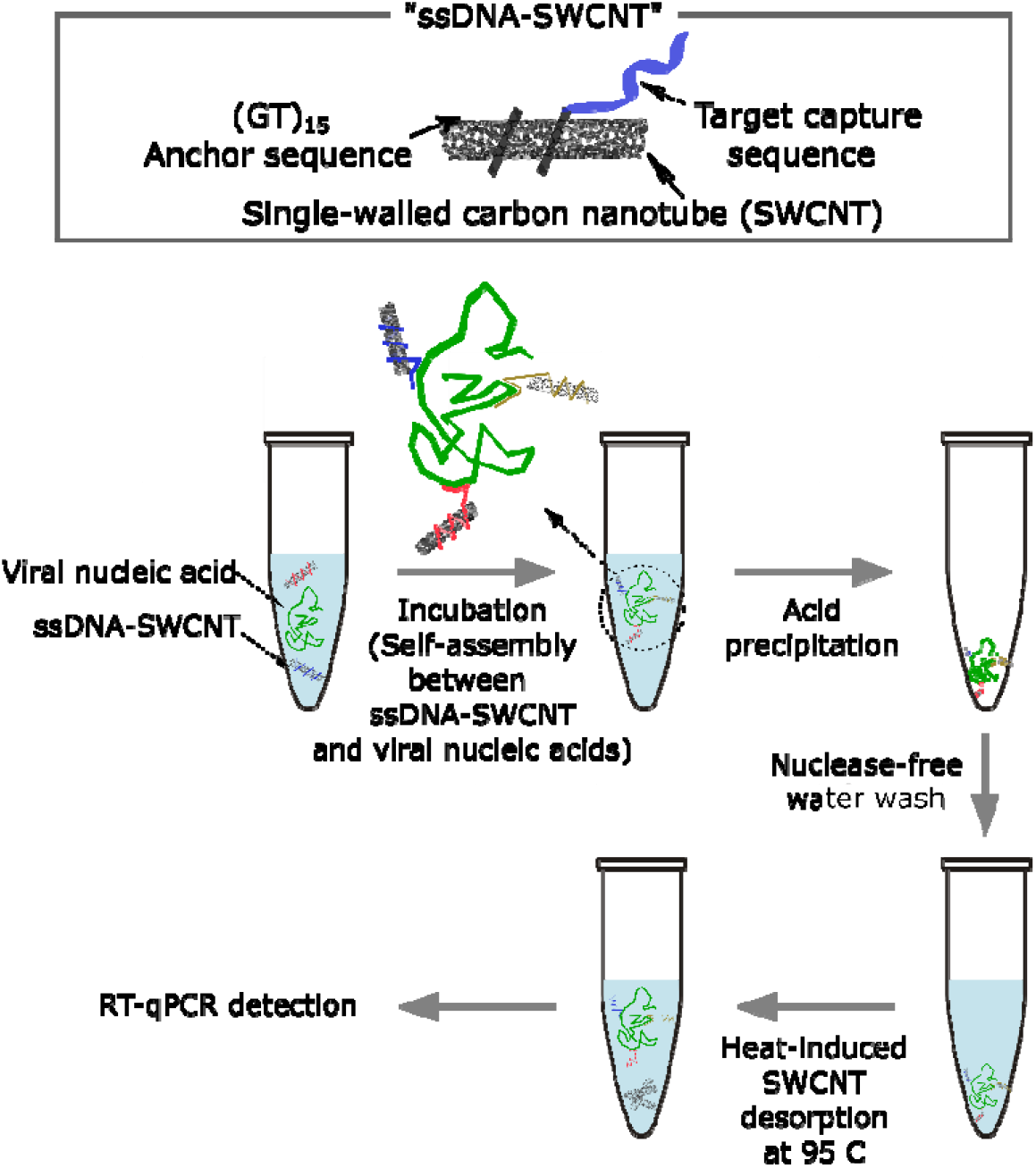
Schematic of single-stranded nucleic acid extraction with ssDNA-SWCNT capturing constructs. (1) Target viral nucleic acid is incubated with ssDNA-functionalized SWCNT capturing constructs, which results in the self-assembly of the target nucleic acid on ssDNA-SWCNTs via nucleic acid hybridization. (2) The self-assembled ssDNA-SWCNT and the captured target nucleic acid is precipitated by centrifugation in acidic conditions. (3) The product is purified with nuclease-free water to remove unwanted biomolecules. (4) The product is heated at 95 □ to desorb the target nucleic acids from the SWCNT surface. (5) The extracted target nucleic acids can be amplified by reverse transcription and polymerase chain reaction.

We first optimized the number of unique ssDNA-SWCNT capture sequences to maximize extraction of SARS-CoV-2 viral RNA. 4×10^6^ copies of synthetic SARS-CoV-2 RNA was spiked into a PBS solution to serve as the RNA standard solution. The RNA from this standard solution was extracted by introducing ssDNA-SWCNT constructs with different capture regions, precipitating, recovering, and detecting the recovered RNA by subsequent RT-qPCR with the corresponding SARS-CoV-2 primer/probe set (Table S2). When four unique ssDNA-SWCNT constructs (Target 1 - Target 4 DNA sequences) were used, 64±8% of the SARS-CoV-2 viral RNA was extracted from the standard solution (Figure 2A). The extraction efficiency was improved to 89±7% when ten unique ssDNA-SWCNT constructs (Target 1 - Target 10 DNA sequences) were used. Conversely, only 13±4% of the viral RNA was recovered in the absence of capturing ssDNA-SWCNT constructs. We hypothesize that the increase in SARS-CoV-2 viral RNA extraction efficiency with an increased number of unique ssDNA-SWCNT capture sequences is due to the ability of the viral RNA to hybridize with more than one ssDNA-SWCNT construct. Therefore, when the number of unique capture sequences increases, the viral RNA bound to multiple ssDNA-SWCNT constructs can be precipitated more easily during the acid precipitation step of the extraction.

**Figure 2.**
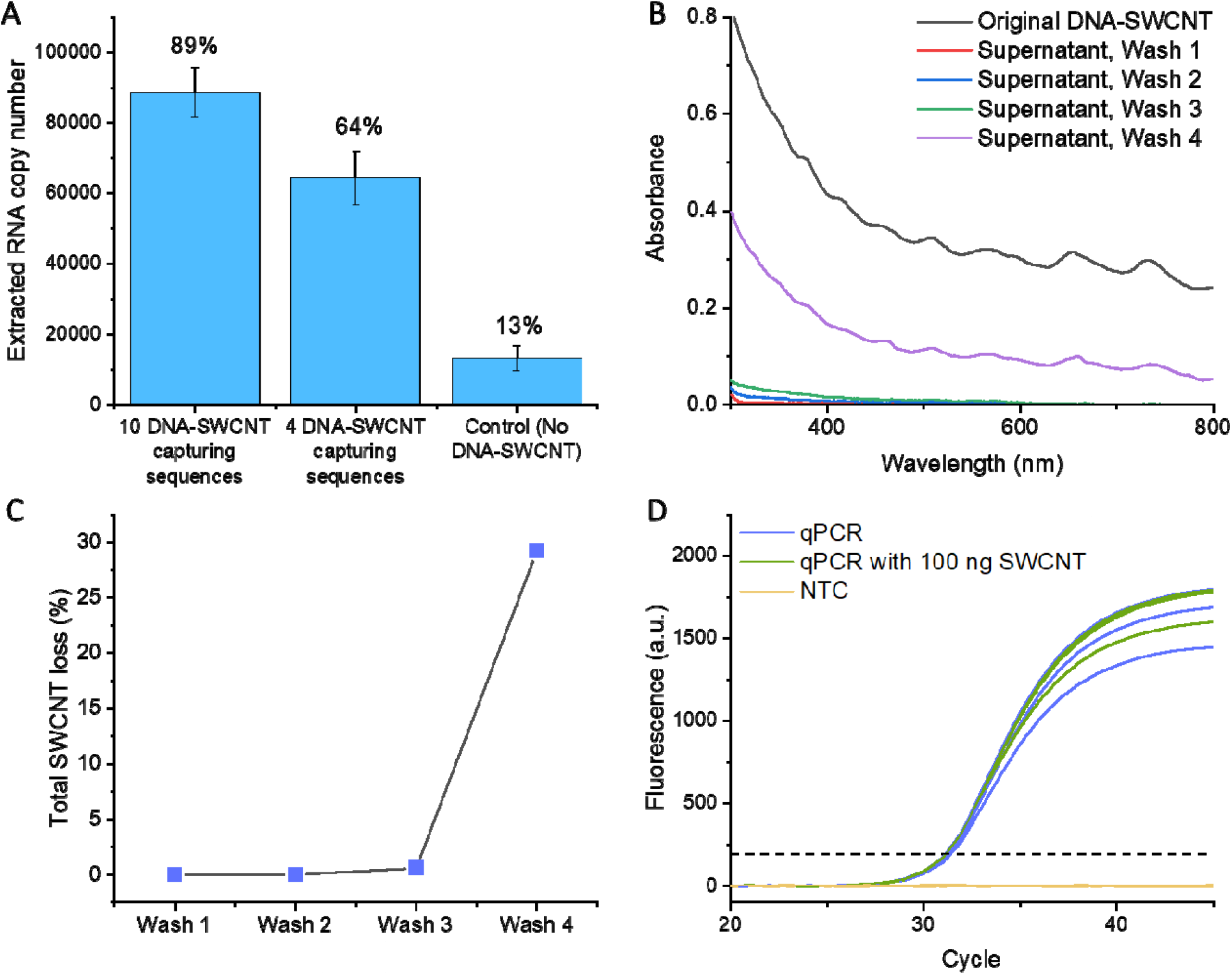
Optimization of SARS-CoV-2 RNA extraction protocol with ssDNA-SWCNT constructs. (A) RNA extraction efficiency is dependent on the number of unique ssDNA capture sequences used to sequester the target viral RNA. Ten or four DNA-SWCNT capturing sequences denotes Targets # 1-10 or Targets # 1-4, respectively, from Table 1. As a control, the extraction protocol is followed without ssDNA-SWCNT. Error bars are standard deviation from n = 3 independent trials. (B-C) The loss of (GT)_15_-SWCNT from the precipitated pellet in acidic conditions after multiple washing steps. (B) Absorption spectra of (GT)_15_-SWCNT in the supernatant after each wash step, and (C) total loss of SWCNT calculated from the absorbance of SWCNT at 632 nm, as a function of the number of wash steps. (D) RT-qPCR compatibility of (GT)_15_-SWCNT. RT-qPCR detection of SARS-CoV-2 RNA is unaffected in the presence of 100 ng (GT)_15_-SWCNT in the reaction tube (n=3). NTC = non template control.

To optimize extraction efficiency at the acid precipitation step, we quantified the amount of ssDNA-SWCNT found in the solution supernatant as a function of the number of pellet washes, seeking to maximize the number of washes while minimizing the disturbance of ssDNA-SWCNT from the pellet into the supernatant. To this end, we used (GT)_15_ ssDNA-SWCNT as a model case, and quantified the SWCNT concentration in the supernatant by measuring the absorbance of SWCNT at 632 nm (Figure 2B and 2C). Following addition of HCl and centrifugation, (GT)_15_-SWCNT were precipitated, and no trace of (GT)_15_-SWCNT was observed in the supernatant. Next, the absorption spectra of the supernatant was monitored following serial supernatant decantation with distilled water. Up to the second water wash, no (GT)_15_-SWCNT were measured in the supernatant, suggesting that two water washes following acid precipitation of ssDNA-SWCNTs is optimal for the acid wash step. Conversely, 0.59% and 29% of (GT)_15_-SWCNT were lost from the pellet and measured in the supernatant following the third and fourth washes, respectively, as the solution became less acidic due to consecutive dilutions. Therefore, for our downstream experiments testing SARS-CoV-2 viral RNA extraction, precipitates were twice-washed during the acid precipitation step to minimize the loss of viral RNA but maximize the purity of the recovered nucleic acids.

Prior work has suggested that carbon nanotubes can adversely affect the efficacy of PCR^16, 17^. Therefore, we also investigated whether any remaining SWCNT in the final extracted RNA product hinders the subsequent RT-qPCR assay. The standard solution spiked with synthetic SARS-CoV-2 RNA was subject to RT-qPCR to detect the viral RNA, with and without 100 ng of (GT)_15_-SWCNT in the reaction tube, which is equal to the maximum amount of SWCNT used for a single extraction step. The PCR amplification curve revealed only marginal interference of SWCNT constructs on quantitation cycle (C_q_) values and curve shapes (Figure 2D), and this result confirms the compatibility of SWCNT for downstream RT-qPCR analysis, in the unlikely event that any were to remain in the supernatant following the acid precipitation and pellet wash step.

The sensitivity of RT-qPCR depends on the efficiency with which the target viral RNA can be extracted. In the case of SARS-CoV-2, the highly variable viral load between individuals motivates extraction of RNA from patient samples (nasal swabs, saliva samples) with maximal efficiency. Therefore, the synthetic SARS-CoV-2 RNA extraction efficiency of our protocol (Figure 1) was compared to that of a commercial viral RNA/DNA extraction kit (PureLink® Viral RNA/DNA Mini Kit, Invitrogen) which uses silica substrates for viral RNA/DNA extraction^18^. To evaluate the extraction efficiency in an inhibitor-free solution, the standard RNA template solution was prepared as described above, by spiking PBS with 2×10^6^ copies synthetic SARS-CoV-2 RNA. Standard solutions were ten-fold serially diluted to probe the working range of nucleic acid copy numbers for extraction. The original spiked RNA samples were also amplified by RT-qPCR without any extraction steps to calculate the baseline RNA recovery yield (Figure 3C). RT-qPCR amplification plots and C_q_ values of RNA extracted from our method vs. the PureLink method are shown in Figure 3. Our SWCNT-based extraction protocol recovered □100% of the spiked RNA across three logs of spiked RNA concentration ranging from 2×10^3^ to 2×10^6^ RNA copies per 0.2 mL with a correlation coefficient (R^2^) of 0.9917 and an amplification efficiency (*E*) of 95%. Conversely, the PureLink commercial extraction kit showed a significantly lower recovery yield of ∼20% with R^2^ of 0.9997 and *E* of 95%. Both extraction protocols can be completed in an hour.

**Figure 3.**
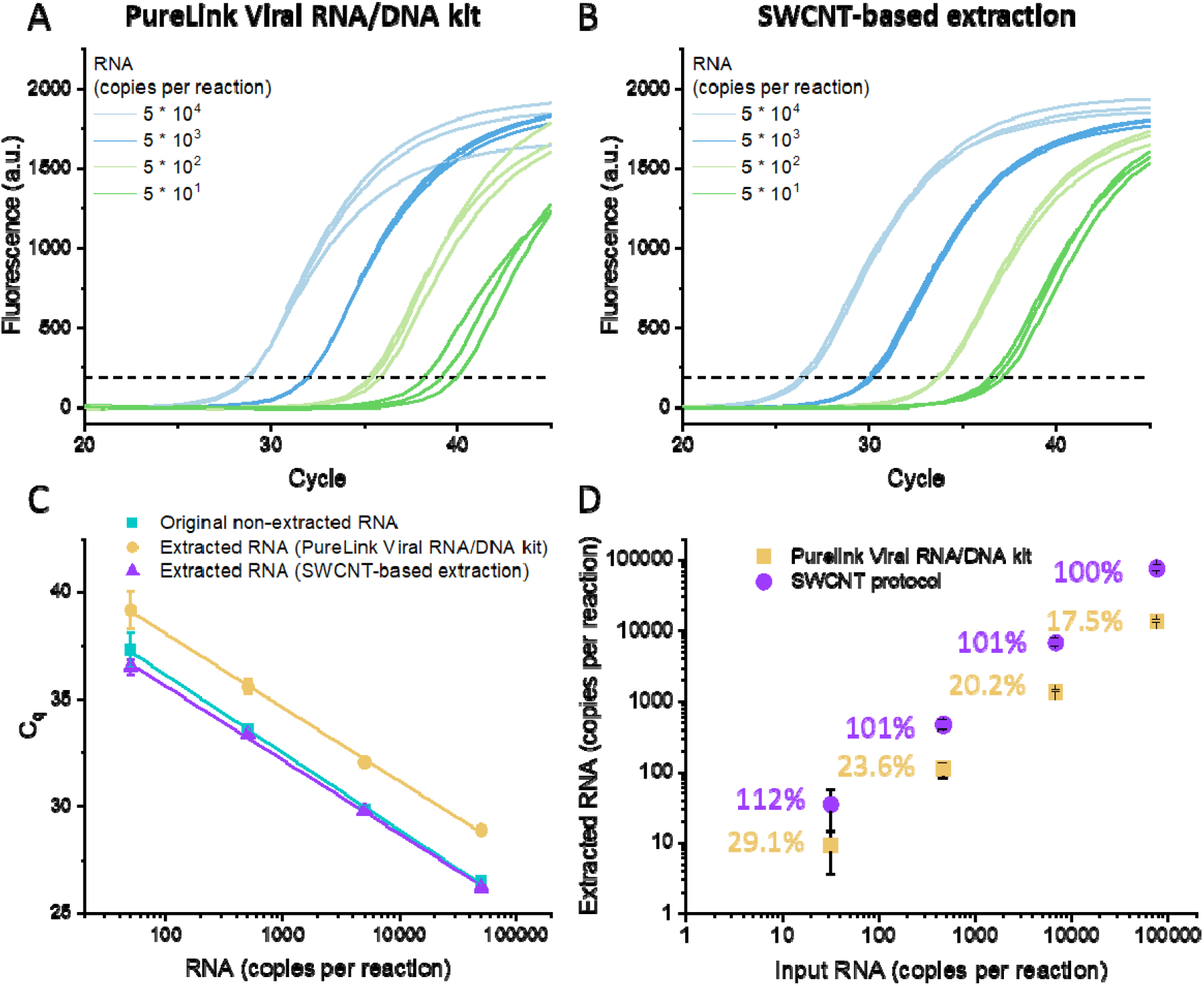
Synthetic SARS-CoV-2 genomic RNA extraction from PBS. RT-qPCR amplification of (A) SARS-CoV-2 RNA extracted by a commercial PureLink Viral RNA/DNA extraction kit, and (B) SARS-CoV-2 RNA extracted by ssDNA-SWCNT-based protocol. RNA templates were serially diluted ten-fold across three orders of magnitude. (C) Comparison of RT-qPCR amplification of original non-extracted RNA (blue), RNA extracted with ssDNA-SWCNT based protocol (purple), and RNA extracted with a commercial PureLink Viral RNA/DNA kit (yellow), where lower C_q_ values represent better extraction of RNA. (D) Extraction efficiency of RNA in PBS with the ssDNA-SWCNT based protocol (purple) and PureLink Viral RNA/DNA kit (yellow). Error bars are standard deviation from n = 3 independent trials.

A major bottleneck for RT-qPCR based detection of viral nucleic acids is the necessity to purify a bio-specimen prior to PCR detection. Therefore, we sought to compare the extraction efficiency of synthetic SARS-CoV-2 RNA from complex human biofluids with our SWCNT-based protocol versus a commercial PureLink kit. We therefore tested the extraction of SARS-CoV-2 RNA from a 50% (v/v) solution of human saliva in PBS. RT-qPCR amplification and C_q_ values were obtained as detailed above. Our results show that SWCNT-based extraction protocol enables recovery of >50% of the SARS-CoV-2 synthetic RNA with R^2^ of 0.9891 and *E* of 99% across three orders of magnitude of concentration from 2_×_10^3^ to 2_×_10^6^ RNA copies per 0.2 mL. In comparison, the PureLink column-based kit showed a similar recovery yield of >60% with R^2^ of 0.9917 and *E* of 87% (Figure 4). Notably, the PureLink kit relies on supply-chain limited reagents and requires 15 steps with four tubes to enable extraction of RNA, whereas, SWCNT-based viral RNA extraction can be done as a ‘one-pot’ protocol in a single tube. Therefore, viral RNA can be directly extracted and PCR-quantified from crude human saliva biofluids with our SWCNT-based protocol, at efficiencies comparable to RNA extraction with commercial DNA/RNA extraction kits.

**Figure 4.**
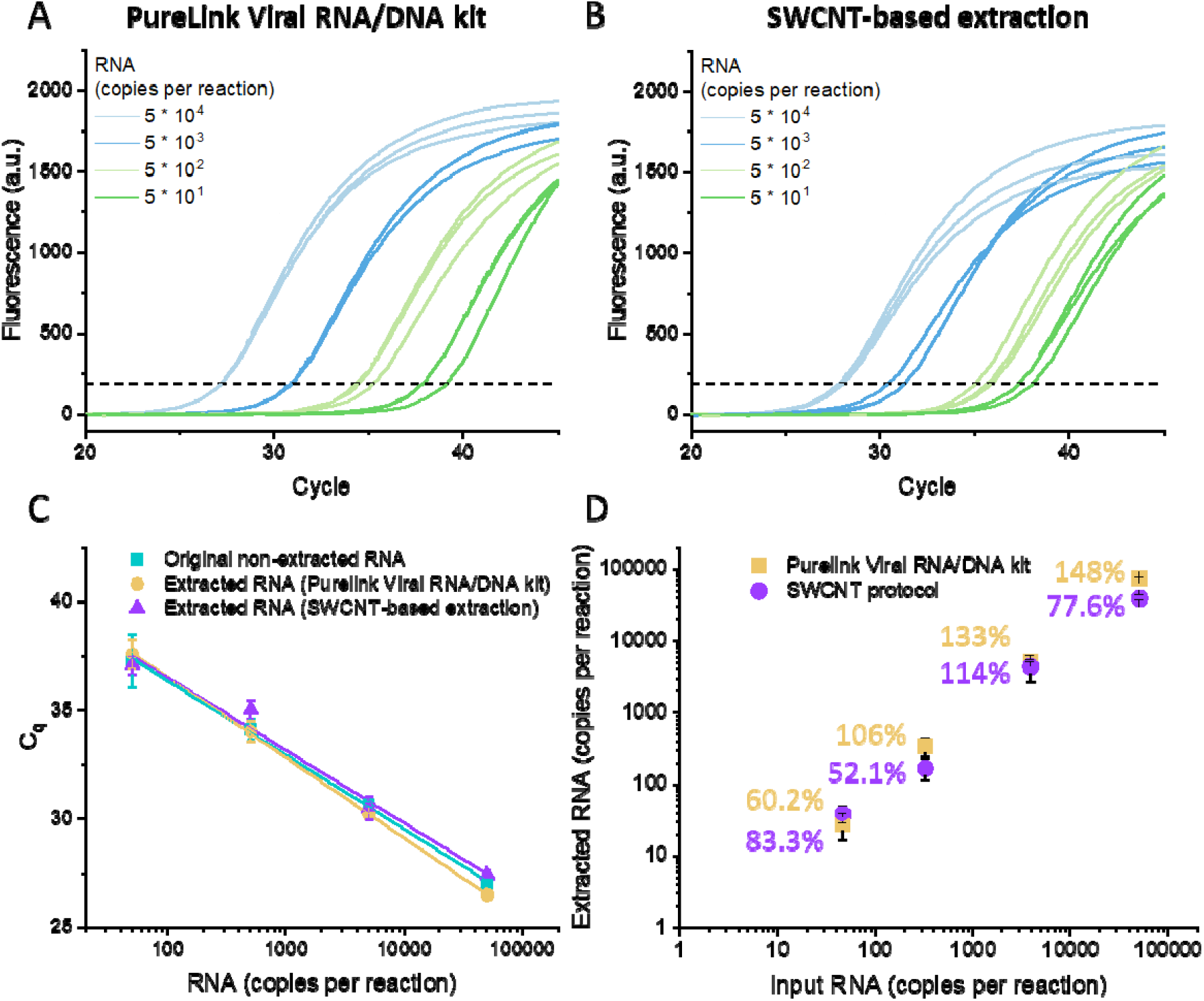
Synthetic SARS-CoV-2 genomic RNA extraction from 50% human saliva. RT-qPCR amplification of (A) SARS-CoV-2 RNA extracted by a commercial PureLink Viral RNA/DNA extraction kit, and (B) SARS-CoV-2 RNA extracted by ssDNA-SWCNT-based protocol. RNA was extracted from 50% human saliva spiked with synthetic SARS-CoV-2 genomic RNA. RNA templates were serially diluted ten-fold across three orders of magnitude. (C) Comparison of RT-qPCR amplification of original non-extracted RNA (blue), RNA extracted with ssDNA-SWCNT-based protocol (purple), and RNA extracted with a commercial PureLink Viral RNA/DNA kit (yellow). (D) Extraction efficiency of RNA from 50% human saliva with the ssDNA-SWCNT based protocol (purple) and PureLink Viral RNA/DNA kit (yellow). Error bars are standard deviation from n = 3 independent trials.

## Discussion

The ability to first extract and next quantify viral nucleic acids from patients forms the foundation of testing and tracking viral infections, such as that for the current SARS-CoV-2 pandemic. Increasing the extraction yield of viral nucleic acids, to do so from crude patient biofluids, and with materials and reagents orthogonal to those in commercial DNA/RNA extraction kits could greatly increase our testing capacity and could be seamlessly applied to all other DNA or RNA detection workflows. In this study, we developed a protocol for extraction of single-stranded viral nucleic acids such as SARS-CoV-2 RNA, for subsequent quantification by RT-qPCR. ssDNA-functionalized SWCNT suspensions with DNA-based ‘capture’ sequences enabled high sensitivity extraction of the SARS-CoV-2 RNA from both PBS and human saliva biofluids. To bind the viral nucleic acid to substrates, our SWCNT-based extraction protocol makes use of ssDNA sequences on SWCNT, which contain sequences complementary to specific regions in the SARS-CoV-2 viral genome, serving as capturing ligands. We demonstrate that our SWCNT-based viral RNA extraction protocol enables higher RNA extraction efficiency in PBS (∼100% extraction efficiency) as compared to a commercial silica-matrix column PureLink kit (∼20% extraction efficiency). Furthermore, we show that SWCNT-based RNA extraction enables us to bypass the lengthy protocols associated with nucleic acid cleanup from patient biofluids by simply mixing ssDNA-SWCNT with human saliva, for facile recovery of viral nucleic acids. Our SWCNT-based extraction protocol combined with SARS-CoV-2 CDC RUO kit represented the limit of quantification (LoQ) of 6.4 copies/μL in original PBS buffer, and LoQ of 9.2 copies/μL in original 50% human saliva.

The distinct advantages of our SWCNT-based protocol are in increasing the sensitivity of viral RNA detection, and in enabling RT-qPCR based viral RNA quantification without silica-column based purification of patient saliva samples. ssDNA-SWCNTs are produced in a ‘chemistry-free’ system *via* sonication-based conjugation of capturing ssDNA to the SWCNT, greatly reducing the cost and time to prepare this nucleic acid extraction system. Furthermore, the approach described herein requires only small quantities of SWCNT and HCl in a one-pot system. As such, our protocol enables purification of viral nucleic acids directly from patient biofluid, avoiding the need for, and time to operate, silica-matrix columns that require numerous binding, washing, and elution reagents and steps. Lastly, the modularity of our approach enables the user to choose any specific capturing ssDNA sequences for attachment to SWCNT, potentially enabling detection of different viral nucleic acids, including ssDNA-genome-based viruses, and positive strand and negative strand RNA viruses.^19, 20^

Taken together, our results demonstrate that ssDNA-SWCNT based extraction of viral nucleic acids is a modular approach that could serve as a low-cost and rapid method to extract target nucleic acids from patient biofluids. The manufacturing burden and cost of ssDNA-SWCNTs are significantly lower than those associated with current viral nucleic acid extraction kits and protocols, enabling this technology to be potentially implemented in any laboratory with modularity that enables researchers to rapidly substitute their desired capture sequences on the SWCNT for detection of other viral or diagnostic nucleic acids.

Pertinent to the current pandemic, this SWCNT-based extraction protocol can alleviate the supply shortages and unprecedented demands for RT-qPCR testing due to its manufacturing simplicity. Lastly, easy substitution of other capture ssDNA on ssDNA-SWCNTs could find applications in detection of other viral infections such as influenza, or applications in routine clinical diagnostics such as genetic disorder testing using cell-free DNA.

## Methods

### A. Preparation of ssDNA-SWCNT constructs with capturing target nucleic acids

For SARS-CoV-2 RNA extraction, 10 unique single-stranded DNA sequences (ssDNAs, purchased from Integrated DNA technologies) were used as viral RNA ‘capture’ sequences, each contain ing 18-mer DNA sequences complementary to different parts of the SARS-CoV-2 viral RNA genome, and (GT)_15_ ‘anchor’ sequences for adsorption to the SWCNT surface. RNA sequences from SARS-CoV-2 (variant MT007544.1) were used to design the capture ssDNA sequences. Of note, the complementary sequence region can be changed according to the desired target nucleic acid sequences to be recovered. Full ssDNA sequences from Target 1 to Target 10 are listed in Table 1. For each ssDNA, ssDNA-functionalized SWCNT constructs were generated with the following protocol: 1 mg of HiPCo SWCNT (small diameter HiPCo SWCNT, NanoIntegris) was added to 0.9 mL of phosphate-buffered saline (PBS), and the solution was mixed with 100 µL of 1 mM ssDNA. The resulting mixture was bath-sonicated for 2 min and probe-tip sonicated for 10 min at 5 W power in an ice bath. After sonication, the ssDNA-SWCNT suspension was centrifuged for 30 mi n at 16,100 g to precipitate unsuspended SWCNT, and the supernatant containing colloidally sus pended ssDNA-SWCNT was collected. The supernatant was spin-filtered with a 100 kDa MWCO centrifugal filter at 6000 rpm for 5 min with DNase free water to remove unbound ssDNA, and th e purified solution at the top of the filter was collected. This spin filtration to remove unbound ssDNA was repeated three times. The ssDNA-SWCNT suspension was diluted with PBS buffer and stored at 4 □C until further use. The concentration of the ssDNA-SWCNT suspension was calculated by measuring absorbance at 632 nm with an extinction coefficient for SWCNT of 0.036 (mg/ L)^-1^ cm^-1^.^21^

### B. Synthetic SARS-CoV-2 viral RNA extraction by ssDNA-SWCNT constructs

10 unique SARS-CoV-2 RNA-capturing ssDNA-SWCNT suspensions were mixed to make a 1 mg/L ssDNA-SWCNT solution for each unique sequence of ssDNA-SWCNT. The concentration of total ssDNA-SWCNT was 10 mg/L. For the extraction of target RNA from PBS buffer, 190 μL o f a known amount of RNA (synthetic SARS-CoV-2 RNA Control 1 (MT007544.1), Twist Bioscience) in PBS buffer was mixed with 10 μL of as-prepared total 10 mg/L ssDNA-SWCNT mixture, and incubated for 10 min at room temperature to enable target RNA capture by ssDNA-SWCNT con structs. Next, 5 μL of 5 M hydrochloric acid was added to precipitate the ssDNA-SWCNT constructs that had bound target RNA. The solution was gently shaken. The solution was centrifuged at 16,000 g for 3 min, and the supernatant was discarded. The precipitates were resuspended in 20 0 μL DNase/RNase free water. The solution was centrifuged again at 16,000 g for 3 min, and the supernatant was discarded. The precipitates were resuspended in 200 μL PBS buffer and briefly sonicated in a bath sonicator (CPX-952-218R, Branson). The mixture solution was incubated at 9 5 °C for 10 min to weaken the adsorption of nucleic acids from the SWCNT surface. The extracte d RNA solution (200 μL) was kept at 4 °C before further use. For downstream RT-qPCR, 5 μL of the extracted RNA solution (2.5% of the total solution) was mixed with the RT-qPCR master mix.

For the extraction of RNA from human saliva, 100 μL of human saliva (pooled normal human saliva, Innovative Research), 85 μL of a known amount of viral RNA in PBS buffer, and 5 μL of 40 U/μL ribonuclease inhibitor (RNaseOUT™Recombinant Ribonuclease Inhibitor, Thermo Fisher) was mixed with 10 μL of as-prepared ssDNA-SWCNT mixture and incubated for 10 min at room temperature. Ribonuclease inhibitor was added to prevent degradation of unprotected RNAs in saliva, which can be replaced with viral lysis buffer in the case of a viral specimen test. The remaining PCR protocol is the same as that for nucleic acid extraction from PBS buffer.

For RNA extraction with a commercial PureLink Viral RNA/DNA Mini Kit (Invitrogen), we followed the protocol provided by manufacturer with the proteinase K and carrier RNA included in the kit.

### C. RT-qPCR protocol

For RT-qPCR, PCR master mix containing GoTaq® 1-Step RT-qPCR System (Promega) master mix, 500 nM of forward and reverse primers, 125 nM of FAM-based probes, and 40 U of RNaseO UT™Recombinant Ribonuclease Inhibitor (Life Technologies, USA) was used. The sequence details of primers and probes are in Table S2. 5 μL of extracted RNA template solution were added in a total volume of 20 μL in triplicate. All reactions were completed in a 96-well plate format (Micr oAmp™Fast Optical 96-Well Reaction Plate with Barcode, 0.1 mL). The RT-qPCR assays were performed under the following conditions: reverse transcription at 45°C for 15 minutes and initial denaturation at 95°C for 2 minutes, 45 cycles of denaturation at 95°C for 3 seconds, and annealing at 55°C for 30 seconds using a standard benchtop real-time thermocycler (CFX96 Touch Real-Time PCR System, BIO-RAD). The C_q_ values and standard deviations obtained were used to calculate the limit of quantification (LoQ). The LoQ was determined with the lowest number of detect able RNA with a coefficient of variation (CV) < 35%, and was estimated by following conventional calculations^22^.

## Data Availability

Data are available from the corresponding author upon request

## Acknowledgements

We acknowledge support of the IGI LGR ERA and Citris/Banatao Seed Funding. We acknowledge support of a Burroughs Wellcome Fund Career Award at the Scientific Interface (CASI) (to M.P.L.), the Simons Foundation (to M.P.L.), a Stanley Fahn PDF Junior Faculty Grant with Award # PF-JFA-1760 (to M.P.L.), a Beckman Foundation Young Investigator Award (to M.P.L.), a CZI investigator award (to M.P.L), a Sloan Foundation Award (to M.P.L.), a Moore Foundation Award (to M.P.L.), a Cisco Research Center grant (to M.P.L), and a DARPA Young Investigator Award (to M.P.L.). M.P.L. is a Chan Zuckerberg Biohub investigator. Work at the Molecular Foundry was supported by the Office of Science, Office of Basic Energy Sciences, of the U.S. Department of Energy under Contract No. DE-AC02-05CH11231. R.L.P., F.L., and D.Y. acknowledge the support of NSF Graduate Research Fellowships (NSF DGE 1752814).

## Author Contributions

S.J. and M.P.L. conceived the idea and designed experiments. S.J. and E.G.G. performed the experiments and characterized the extraction protocols. All authors discussed the results and wrote the manuscript.

## References

1. Bruce, E.A. et al. Direct RT-qPCR detection of SARS-CoV-2 RNA from patient nasopharyngeal swabs without an RNA extraction step. PLoS Biol. 18, e3000896 (2020).

2. Amen, A.M. et al. Blueprint for a pop-up SARS-CoV-2 testing lab. Nat. Biotechnol. 38, 791–797 (2020).

3. Esbin, M.N. et al. Overcoming the bottleneck to widespread testing: a rapid review of nucleic acid testing approaches for COVID-19 detection. RNA 26, 771–783 (2020).

4. Smyrlaki, I. et al. Massive and rapid COVID-19 testing is feasible by extraction-free SARS-CoV-2 RT-PCR. Nat. Commun. 11, 4812 (2020).

5. Won, J. et al. Development of a Laboratory-safe and Low-cost Detection Protocol for SARS-CoV-2 of the Coronavirus Disease 2019 (COVID-19). Exp. Neurobiol. 29, 107–119 (2020).

6. Sentmanat, M., Kouranova, E. & Cui, X. One-step RNA extraction for RT-qPCR detection of 2019-nCoV. bioRxiv, 2020.2004.2002.022384 (2020).

7. Ali, N., Rampazzo, R.d.C.P., Costa, A.D.T. & Krieger, M.A. Current Nucleic Acid Extraction Methods and Their Implications to Point-of-Care Diagnostics. Biomed. Res. Int. 2017, 9306564–9306564 (2017).

8. Wang, J. et al. Fast DNA Extraction with Polyacrylamide Microspheres for Polymerase Chain Reaction Detection. ACS Omega 5, 13829–13839 (2020).

9. Kolluri, N. et al. SNAPflex: a paper-and-plastic device for instrument-free RNA and DNA extraction from whole blood. Lab Chip (2020).

10. Berensmeier, S. Magnetic particles for the separation and purification of nucleic acids. Appl. Microbiol. Biotechnol. 73, 495–504 (2006).

11. Gopal, H. et al. Oligonucleotide Based Magnetic Bead Capture of Onchocerca volvulus DNA for PCR Pool Screening of Vector Black Flies. PLoS Negl. Trop. Dis. 6, e1712 (2012).

12. Hei, A.-L. & Cai, J.-P. Development of a Method for Concentrating and Purifying SARS Coronavirus RNA by a Magnetic Bead Capture System. DNA Cell Biol. 24, 479–484 (2005).

13. Chio, L. et al. Electrostatic Assemblies of Single-Walled Carbon Nanotubes and Sequence-Tunable Peptoid Polymers Detect a Lectin Protein and Its Target Sugars. Nano Lett. (2019).

14. Gigliotti, B., Sakizzie, B., Bethune, D.S., Shelby, R.M. & Cha, J.N. Sequence-Independent Helical Wrapping of Single-Walled Carbon Nanotubes by Long Genomic DNA. Nano Lett. 6, 159–164 (2006).

15. Harvey, J.D. et al. A carbon nanotube reporter of microRNA hybridization events in vivo. Nat. Biomed. Eng. 1, 0041 (2017).

16. Zhang, Z., Shen, C., Wang, M., Han, H. & Cao, X. Aqueous suspension of carbon nanotubes enhances the specificity of long PCR. Biotechniques 44, 537–545 (2008).

17. Williams, R.M., Nayeem, S., Dolash, B.D. & Sooter, L.J. The Effect of DNA-Dispersed Single-Walled Carbon Nanotubes on the Polymerase Chain Reaction. PLoS ONE 9, e94117 (2014).

18. Sathiamoorthy, S., Malott, R.J., Gisonni-Lex, L. & Ng, S.H.S. Selection and evaluation of an efficient method for the recovery of viral nucleic acids from complex biologicals. NPJ Vaccines 3, 31 (2018).

19. Tran, T.L. et al. Detection of influenza A virus using carbon nanotubes field effect transistor based DNA sensor. Physica E Low Dimens. Syst. Nanostruct. 93, 83–86 (2017).

20. Tam, P.D., Van Hieu, N., Chien, N.D., Le, A.-T. & Anh Tuan, M. DNA sensor development based on multi-wall carbon nanotubes for label-free influenza virus (type A) detection. J. Immunol. Methods 350, 118–124 (2009).

21. Zhang, J. et al. Molecular recognition using corona phase complexes made of synthetic polymers adsorbed on carbon nanotubes. Nat. Nanotechnol. 8, 959–968 (2013).

22. Forootan, A. et al. Methods to determine limit of detection and limit of quantification in quantitative real-time PCR (qPCR). Biomol. Detect. Quantif. 12, 1–6 (2017).

