## Supplemental Information for "Extraction of Viral Nucleic Acids with Carbon Nanotubes Increases SARS-CoV-2 RT-qPCR Detection Sensitivity"

| Name | Target Sequence in SARS-CoV-2 | Target Region in SARS-CoV-2 |
|---|---|---|
| Target1 | AAAAAAAAAAAAAAAAAAAAA (poly A tail) | 29861-29893 |
| Target2 | GCUGCUGAGGCUUCUAAG | 29024-29041 |
| Target3 | UUACAAACAUUGGCCGCA | 29164-29181 |
| Target4 | AAAUUUCAAAGAUCAAGU | 29304-29322 |
| Target5 | ACUGUGACUCUUCUUCCU | 29444-29461 |
| Target6 | UUUUCGCUUUUCCGUUUA | 29574-29591 |
| Target7 | GAACUUCUCCUGCUAGAA | 28884-28901 |
| Target8 | GUGCUACAACUUCCUCAA | 28745-28762 |
| Target9 | ACUACCUAGGAACUGGGC | 28605-28622 |
| Target10 | UAAAUUCCCUCGAGGACA | 28465-28482 |

**Table S1.** Sequences targeted in the SARS-CoV-2 viral genome (variant MT007544.1) with 10 target DNA sequences. Base location in the genome for each target is shown. All the target sequences, except Target 1, aim to bind N gene region in the SARS-CoV-2 RNA genome.

| | | |
|---|---|---|
| SARSCoV-2_N1 | SARS-CoV-2_N1-F | GAC CCC AAA ATC AGC GAA AT |
| | SARS-CoV-2_N1-R | TCT GGT TAC TGC CAG TTG AAT CTG |
| | SARS-CoV-2_N1Probe | FAM-ACC CCG CAT TAC GTT TGG TGG ACG BHQ1 |

**Table S2**. Primer and probes used for RT-qPCR detection of SARS-CoV-2 viral RNA. All reagents come from the 2019-nCoV RUO Kit (Integrated DNA technologies).
